# Respiratory viral infections by Non-influenza viruses are associated with more adverse clinical outcome in patients with underlying liver disease: a single centre laboratory based study

**DOI:** 10.1101/2020.08.01.20166330

**Authors:** Ekta Gupta, Abhishek Padhi, Kavita Agarwal, Krithiga Ramachandran, Reshu Agarwal, Samba Siva Rao Pasupuleti, Debajyoti Bhattacharyya, Rakhi Maiwall, Shiv Kumar Sarin

**Author notes:** Corresponding Author Dr Ekta Gupta Professor and Head, Department of Clinical Virology, Institute of Liver and Biliary Sciences, New Delhi, Email ID.

## Abstract

**Background:** Respiratory viral infections are an important cause of acute respiratory tract infections. They are caused by both Influenza and non influenza viruses. Respiratory viral infections are known to be associated with severe clinical outcome especially in the critically ill. A constant surveillance is needed for early etiological identification which can help in timely and appropriate management and will further help in prevention of indiscriminate use of antibiotics in patients with viral etiology.

**Methods:** In this retrospective study, clinical records of all adult liver disease patients with clinically confirmed ARI, whose request for respiratory viral testing were received in the virology laboratory during September 2016 - March 2019 were reviewed. Respiratory viruses were identified by real time PCR on FilmArray 2.0 instrument (BioFire Diagnostics, Utah, USA) using Respiratory panel as per the manufacturer’s instructions.

**Results:** Of the 603 patients of liver disease with clinically confirmed influenza like illness, over all incidence of respiratory viral infection was 24.3% (n= 147). Infections by non-influenza viruses (87, 59.1%) were more than influenza group of viruses. Mortality was higher in non influenza group (43, 49.4%) as compared to influenza (24, 40%) [p=0.015] being maximum in Rhinovirus, 22 (32.8%). Two peaks were observed in both influenza and non influenza groups, first in the months of January and February and the other one in August and October.

**Conclusion:** With the emergence of SARS-CoV-2 it has now become imperative for a constant surveillance of the non influenza viruses for early etiological identification of the respiratory viral infection for proper and timely management in the critically ill.

**Highlights:**

- Patients with liver cirrhosis having Respiratory viral infections have a poor outcome in terms of morbidity and mortality.
- Mortality associated with non influenza viruses (NIV) is more as compared to influenza virus infections.
- COVID-19 pandemic and higher mortality in NIVs warrants a constant monitoring of respiratory viral infections.

## BACKGROUND

Acute respiratory tract infections (ARI) are usually caused by viruses known as respiratory viral infections (RVI). In adult patients with hospital-acquired pneumonia (HAP) RVI is usually seen in 10-23% of the cases [1]. Disease severity and clinical outcome in RVI varies by the type of virus and underlying co-morbidity [2]. Influenza virus (IVI) infections predominate but other Non-Influenza Viruses (NIV) should also be identified. Number of viruses are included in the NIV category like respiratory syncytial virus, para-influenza virus, cytomegalovirus, human corona viruses, bocavirus, human metapneumovirus and adenovirus [1]. Rhinoviruses are often the most common virus in the NIV group [3]. Corona viruses are also associated with severe ARI and the present ongoing pandemic of SARS CoV-2 further emphasizes the need to identify NIV causing pneumonia [4-6]. Advancement in molecular diagnostics and with the easy availability of Polymerase chain reaction (PCR) for most of the respiratory viruses, identification of viral etiology is faster and easier [7]. Early and exact etiological identification in RVI can help in timely and appropriate management and will further help in prevention of indiscriminate use of antibiotics in patients with viral etiology. Timely and appropriate isolation of infected subjects is also important especially in critical care settings [8, 9]. Underlying co-morbid conditions also influence the clinical outcome in pneumonia patients. Liver disease with cirrhosis is considered as an important co-morbid condition. Influenza outcomes in liver disease patients especially that are critically ill have been well characterised and our previous experience has also shown that liver disease patients with Influenza infection are associated with severe clinical outcome [10]. Though NIV are common in adult patients but are studied very less due to limited attention from the clinical interest, lack of availability of diagnostic tools, vaccine or specific anti-viral treatment.

The recent novel corona virus outbreak has shown that there is a need of regular surveillance for NIV.

Therefore, in this single centre hospital based retrospective study, we tried to look at various NIV and compare with IVI in terms of clinical outcome. In this study, three influenza seasons were included to understand the trend of IVI and NIV in Delhi, a city in the northern part of India.

## METHODS

### Study group

In this retrospective study, clinical records of all adult liver disease patients with clinically confirmed ARI, whose request for respiratory viral testing were received in the virology laboratory during September 2016 - March 2019 were reviewed. ARI was defined as per WHO case definition for Influenza like illness as an acute respiratory illness with a measured temperature of ≥ 38 °C and cough, with onset within the past 10 days [11]. Liver disease was considered in patients having abnormal liver function test parameters. Children < 18 years of age, patients with frank immune-suppression, pregnant females, HIV infected and patients with hepato-cellular carcinoma or any other kind of cancer were excluded from this data. Patient’s complete clinical details and follow up investigations were collected from the Hospital information system (HIS).

### Ethics statement

The study was reviewed and approved by the Institutional Review Board, Institute of liver and biliary sciences, New Delhi (ILBS). The informed consent was waived by the Institutional Review Board because the study was based on the retrospective analysis of existing clinical data. Patient records/ information was anonymised and de-identified prior to analysis.

### Respiratory Virus PCR

For respiratory virus testing, combined throat and nasopharyngeal swabs in viral transport media (VTM) were collected and transported maintaining appropriate cold chain (2-8°C) to the virology lab within 4 hours of collection. The swabs along with the VTM were vortexed and about 300µl of the sample was taken up for testing for a panel of 21 different respiratory viruses by real time PCR on FilmArray 2.0 instrument (BioFire Diagnostics, Utah, USA) using Respiratory panel as per the manufacturer’s instructions [12]. The panel included Adenovirus, Corona virus 229E (CoV229E), Corona virus HKU1 (CoVHKU1), Corona virus OC43 (CoVOC43), Corona virus NL63 (CoVNL63), Human Meta-pneumovirus (MPV), Human Rhinovirus/enterovirus (Rhino/entero), Influenza A (Flu A), Influenza A/H1 (Flu A/H1), Influenza A/H1-2009 (Flu A/H1-2009), Influenza A/H3 (Flu A/H3), Influenza B (Flu B), Middle East Respiratory Syndrome Corona virus (MERSCoV), Para-influenza 1 (PIV1), Para-influenza 2 (PIV2), Para-influenza 3 (PIV 3), Para-influenza 4 (PIV 4), Respiratory syncytial virus (RSV). The FilmArray Respiratory Panel (FilmArray RP) is both FDA-approved and CE IVD-marked. The test is performed in a closed system that requires 5 minutes of hands-on time and 65 minutes of instrumentation time. Several comparison studies between FilmArray and other PCR tests for respiratory viruses showed comparable results [13-15].

### Statistical analysis

Results were expressed as mean and standard deviation (SD) and n (%) for qualitative variables. Association of RVI positivity and outcome with patient’s characteristics and biochemical parameters were analyzed using chi-square test/ Fisher’s exact or Wilcoxon rank sum test, as appropriate, for univariate comparison. For multivariable analysis, we used logistic regression model, using RVI positivity (positive/negative) or outcome with mortality (present/absent) as the dependent variable. Co-variables tested in the multivariable model were all variables expected to be associated with dependent variable. Results were expressed as odds ratios (OR) with 95% confidence intervals (CI). A P value of 0.05 or less was considered statistically significant. Statistical analyses were performed using the SPSS software version 25(IBM Inc., Chicago, IL, USA).

## RESULTS

### Baseline characteristics

Six hundred and three patients of liver disease with clinically confirmed ILI were included in the study. In the study population most of them were male (492, 81.6%) and were with a median age of 48 (IQR: 37-59) years **(Table 1)**. Overall incidence of RVI in the study group was 24.3%, n=147. The incidence was significantly more in patients requiring ICU care (64.3%, 388), admitted in various wards (31.3%, 189) than coming to the Out-patient department (OPD) (4.3%,26)(p value: 0.04).

**Table 1:** Baseline characteristics of the study population

| S. No | Variable | Total<br>(n = 603)<br>n (%) | With RVI<br>(n=147)<br>n (%) | Without RVI<br>(n=456)<br>n (%) | p-value |
| --- | --- | --- | --- | --- | --- |
| 1 | Age (in years) |  |  |  | 0.16 |
|  | 18-30 | 82(13.5) | 26 (17.7) | 56 (12.3) |  |
|  | 31-45 | 185(30.6) | 47 (31.9) | 138 (30.2) |  |
|  | 46-60 | 209(34.6) | 50 (34.0) | 159 (34.8) |  |
|  | >60 | 127(21.06) | 24 (16.3) | 103 (22.6) |  |
| 2 | Sex |  |  |  | 0.63 |
|  | Male | 492(81.6) | 118 (80.2) | 374 (82) |  |
|  | Female | 111(18.40) | 29 (19.7) | 82 (17.9) |  |
| 3 | Admission Type |  |  |  | 0.04 |
|  | ICU | 388(64.3) | 101 (68.7) | 287 (62.9) |  |
|  | Ward | 189(31.3) | 36 (24.5) | 153 (33.5) |  |
|  | OPD | 26(4.3) | 10 (6.8) | 16(3.51) |  |
| 4 | Cirrhosis<br>Present<br>Absent | 381(63.1)<br>222(36.8) | 99 (67.3)<br>48 (31.2) | 282 (61.8)<br>174 (38.1) | 0.20 |
| 5 | OUTCOME<br>Expired<br>Stable | 208(34.4)<br>395(65.5) | 77 (52.3)<br>70 (47.6) | 131 (28.7)<br>325 (71.2) | 0.38* |
| 6 | Child–Pugh class | n=381 | n=99(25.9) | n=282(74.0) | 0.006* |
|  | A | 25(6.5) | 13 (13.1) | 13 (4.6) |  |
|  | B | 53(13.9) | 8 (8.0) | 45 (15.9) |  |
|  | C | 303(79.5) | 78 (78.7) | 224 (79.4) |  |
| 7 | MELD | n= 350<br>29.24±8.57 | n=79<br>30.39±8.29 | n=271<br>29.04±8.62 | 0.185 |
| 8 | Total Bilirubin | 9.85±10.13 | 10.34±10.67 | 9.72±9.95 | 0.715 |
| 9 | AST | 253.72±1342.87 | 249.39±1320.31 | 251.88±1338.34 | 0.557 |
| 10 | ALT | 104.08±343.84 | 103.56±395.10 | 103.34±323.07 | 0.094 |
| 11 | ALP | 114.93±94.22 | 112.09±90.550 | 115.51±94.95 | 0.871 |
| 12 | GGT | 73.63±146.367 | 71.65±119.62 | 74.04±152.86 | 0.502 |
| 13 | Urea | 83.63±164.03 | 76.75±53.06 | 86.57±184.52 | 0.853 |
| 14 | Creatinine | 1.88±4.24 | 1.57±1.42 | 1.99±4.76 | 0.534 |
| 15 | Hemoglobin | 8.85±3.68 | 9.22±6.51 | 8.75±2.09 | 0.796 |
| 16 | Platelet count | 101.86±87.50 | 104.46±93.94 | 100.93±85.56 | 0.996 |

Patients with cirrhosis did demonstrate increase incidence of ILI whether due to RVI or without it, but in univariate analysis it was not significant. However multivariate logistic regression model showed higher RVI occurrence in cirrhotic patients as compared to non-cirrhotic patients (OR 2.98 [1.27-7.00], p=0.012).

There was also significant association of ILI occurrence with the Child–Pugh class A to C. Maximum occurrence seen in CHILD C class (79.5%,303) as compared to CHILD B (13.9%,53) and A (6.5%,25), p value : 0.006. This trend was similar in both the groups with RVI and without RVI. Mortality was much higher in the RVI group (52.3%, 77) as compared to overall (34.4%,208) and without RVI (28.7%,131)(p value: 0.38).

### Comparison of RVI cases due to influenza virus infections (IVI) and Non influenza virus infections (NIVI)

Out of 147 patients with RVI, infections by non-influenza viruses (NIV) (87, 59.1%) were more than influenza group of viruses (IVI) including influenza A (H1N1) pdm 2009, Influenza A (H3N2) and Influenza B (60, 40.8%) as depicted in figure 1.

**Figure 1.**
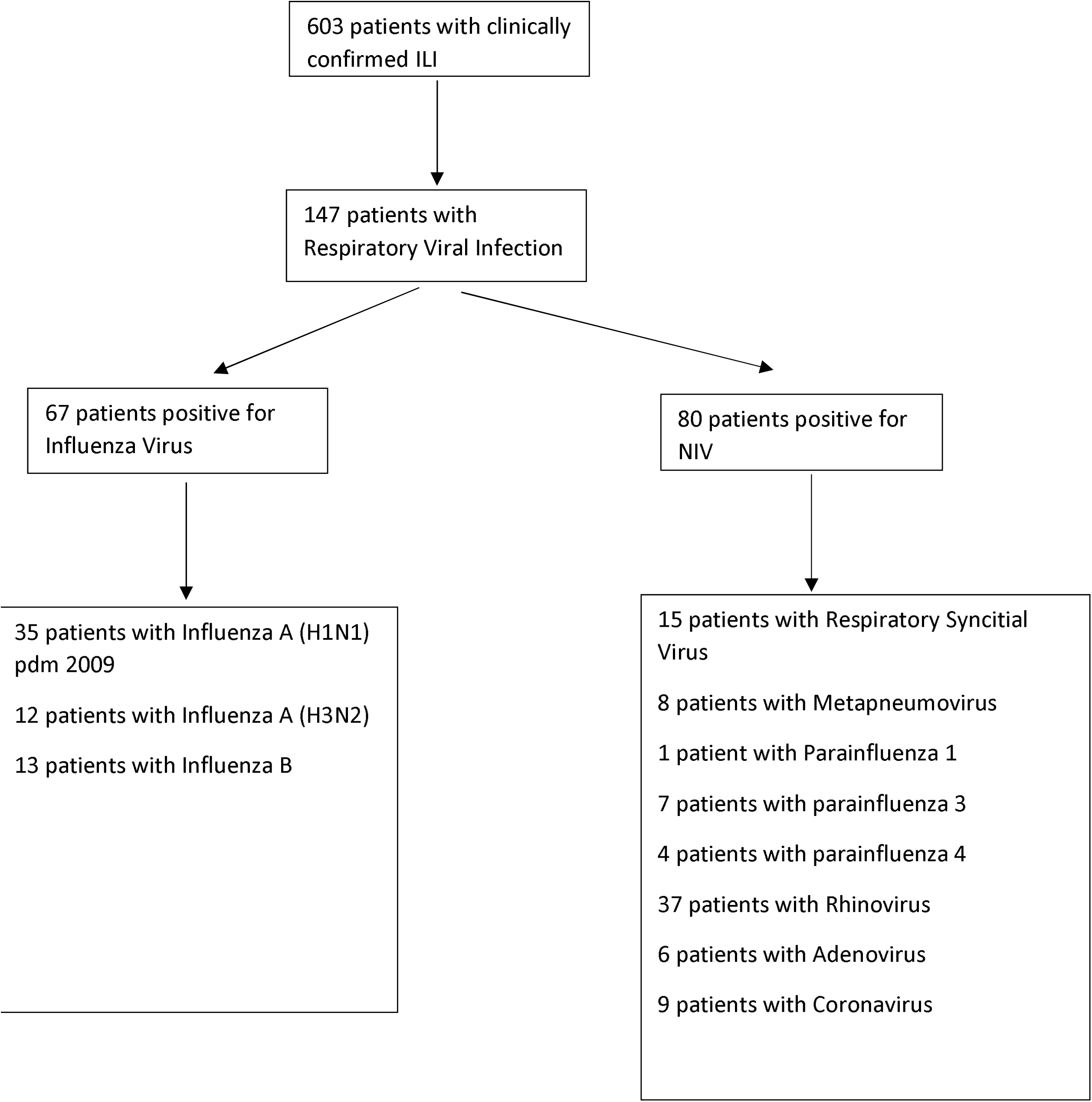
Flowchart of patients hospitalized with influenza-like illness, and viruses detected in respiratory sample

In 8 (5.4%) dual viral infections were also seen, mostly with Rhino/entero group of viruses (Table 2).

**Table 2:** Types of Respiratory Viruses causing infection in the study group.

| Respiratory viruses | n (%) |
| --- | --- |
| Flu A/H1-2009 | 31 (21.1) |
| Flu A /H3 | 12 (8.1) |
| Flu B | 9 (6.1) |
| RSV | 15(10.2) |
| MPV | 8(5.4) |
| PIV 1 | 1 (0.6) |
| PIV3 | 7 (4.7) |
| PIV 4 | 4 (2.7) |
| Rhino/entero | 37 (25.1) |
| Adenovirus | 6 (4.1) |
| Coronavirus* | 9 (6.1) |
| Influenza A (H1N1) pdm 2009+ Rhinovirus | 4(2.7) |
| Influenza A (H1N1) pdm 2009+ Metapneumovirus | 2(1.3) |
| Influenza B+ RSV | 2(1.3) |
| Total | 147(100) |

**Abbreviations:**
Human Meta-pneumovirus (MPV), Human Rhinovirus/enterovirus (Rhino/entero), Influenza A (Flu A), Influenza A/H1-2009 (Flu A/H1-2009), Influenza A/H3 (Flu A/H3), Influenza B (Flu B), Para-influenza 1 (PIV1), Para-influenza 3 (PIV 3), Para-influenza 4 (PIV 4), Respiratory syncytial virus (RSV).
\*Corona virus included : corona virus 229E (CoV229E), corona virus HKU1 (CoVHKU1), corona virus OC43 (CoVOC43), corona virus NL63 (CoVNL63).

In the NIV group, Rhino/ Entero viruses were commonest (41, 27.8%) followed by RSV (17, 11.5%).

Differences among IVI and NIVI groups are presented in Table 3.

**Table 3:**
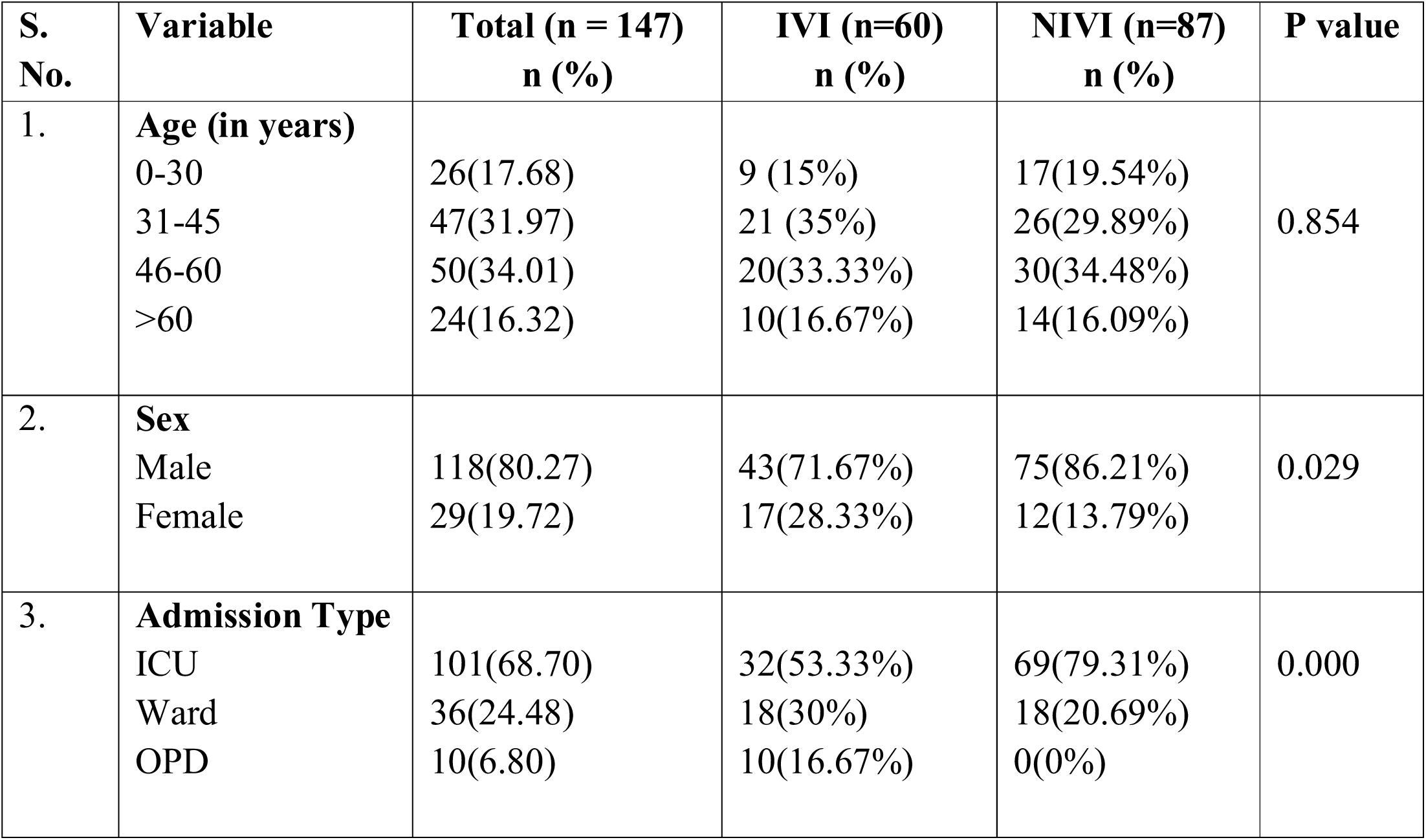
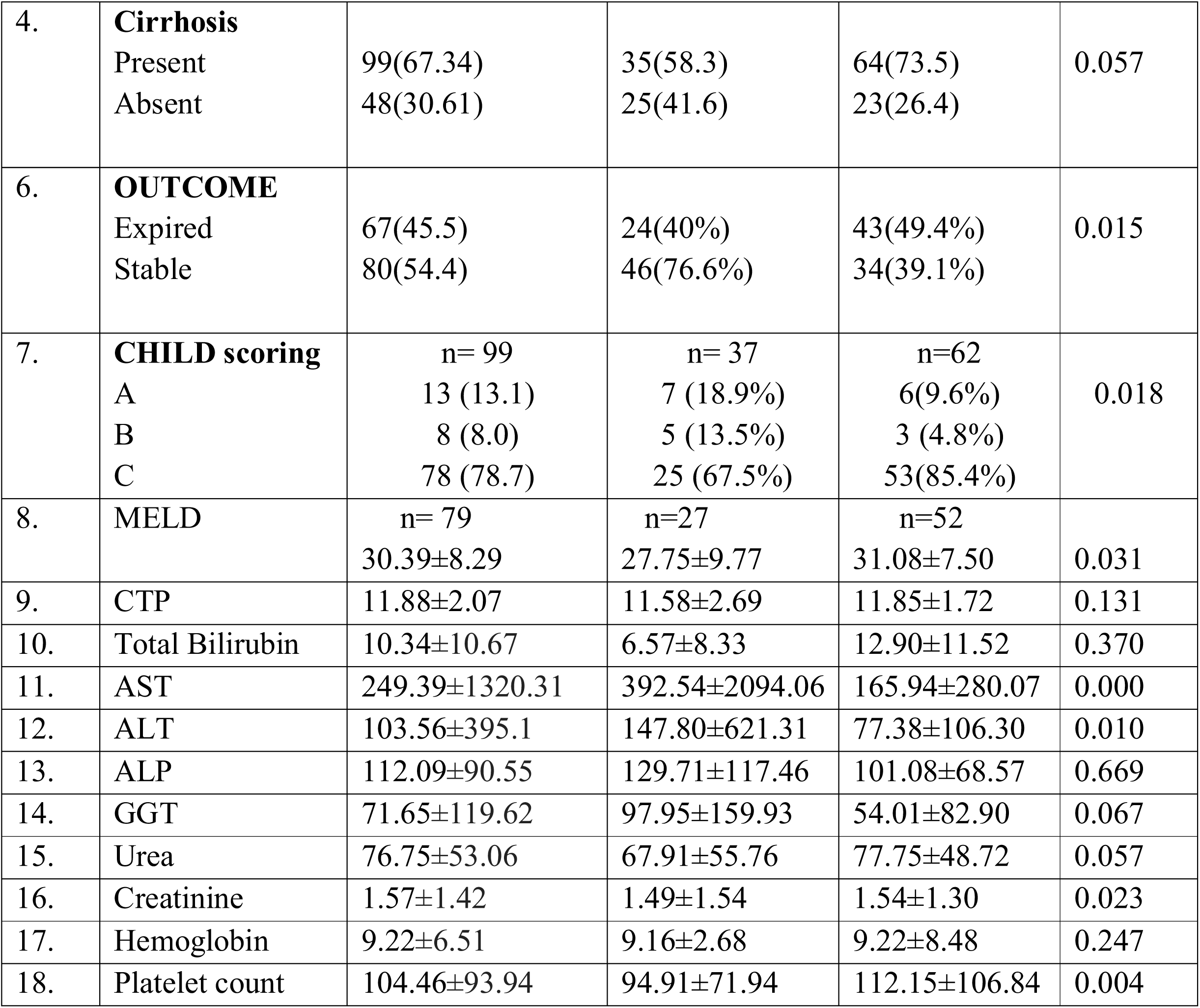
Comparison between Respiratory Viral Infections (RVI) due to Influenza virus infection (IVI) and Non influenza Viral Infections (NIVI).

| S. No. | Variable | Total (n = 147)<br>n (%) | IVI (n=60)<br>n (%) | NIVI (n=87)<br>n (%) | P value |
| --- | --- | --- | --- | --- | --- |
| 1. | **Age (in years)** |  |  |  |  |
|  | 0-30 | 26(17.68) | 9 (15%) | 17(19.54%) | 0.854 |
|  | 31-45 | 47(31.97) | 21 (35%) | 26(29.89%) |  |
|  | 46-60 | 50(34.01) | 20(33.33%) | 30(34.48%) |  |
|  | >60 | 24(16.32) | 10(16.67%) | 14(16.09%) |  |
| 2. | **Sex** |  |  |  |  |
|  | Male | 118(80.27) | 43(71.67%) | 75(86.21%) | 0.029 |
|  | Female | 29(19.72) | 17(28.33%) | 12(13.79%) |  |
| 3. | **Admission Type** |  |  |  |  |
|  | ICU | 101(68.70) | 32(53.33%) | 69(79.31%) | 0.000 |
|  | Ward | 36(24.48) | 18(30%) | 18(20.69%) |  |
|  | OPD | 10(6.80) | 10(16.67%) | 0(0%) |  |
| 4. | **Cirrhosis**<br>Present<br>Absent | 99(67.34)<br>48(30.61) | 35(58.3)<br>25(41.6) | 64(73.5)<br>23(26.4) | 0.057 |
| 6. | **OUTCOME**<br>Expired<br>Stable | 67(45.5)<br>80(54.4) | 24(40%)<br>46(76.6%) | 43(49.4%)<br>34(39.1%) | 0.015 |
| 7. | **CHILD scoring**<br>A<br>B<br>C | n= 99<br>13 (13.1)<br>8 (8.0)<br>78 (78.7) | n= 37<br>7 (18.9%)<br>5 (13.5%)<br>25 (67.5%) | n=62<br>6(9.6%)<br>3 (4.8%)<br>53(85.4%) | 0.018 |
| 8. | MELD | n= 79<br>30.39±8.29 | n=27<br>27.75±9.77 | n=52<br>31.08±7.50 | 0.031 |
| 9. | CTP | 11.88±2.07 | 11.58±2.69 | 11.85±1.72 | 0.131 |
| 10. | Total Bilirubin | 10.34±10.67 | 6.57±8.33 | 12.90±11.52 | 0.370 |
| 11. | AST | 249.39±1320.31 | 392.54±2094.06 | 165.94±280.07 | 0.000 |
| 12. | ALT | 103.56±395.1 | 147.80±621.31 | 77.38±106.30 | 0.010 |
| 13. | ALP | 112.09±90.55 | 129.71±117.46 | 101.08±68.57 | 0.669 |
| 14. | GGT | 71.65±119.62 | 97.95±159.93 | 54.01±82.90 | 0.067 |
| 15. | Urea | 76.75±53.06 | 67.91±55.76 | 77.75±48.72 | 0.057 |
| 16. | Creatinine | 1.57±1.42 | 1.49±1.54 | 1.54±1.30 | 0.023 |
| 17. | Hemoglobin | 9.22±6.51 | 9.16±2.68 | 9.22±8.48 | 0.247 |
| 18. | Platelet count | 104.46±93.94 | 94.91±71.94 | 112.15±106.84 | 0.004 |

Both IVI and NIVI were more frequent in males as compared to females (p=0.029) and in patients who required ICU care (p<0.001).

Differences among the groups were significant in patients with cirrhosis.

Significant differences were seen in the outcome in both the groups, mortality was higher when RVI was due to NIVI than IVI (p=0.015).

Both the groups showed increased association with the CHILD category, being higher in the category C patients. MELD score was higher in the NIVI group (0.031).This could be the reason for more mortality in this group. The liver function tests (ALT,AST) were significantly more deranged in the IVI group than NIVI.

### Respiratory Viral Infections and seasonality

We retrospectively analysed the data from the laboratory archive and the time line included was from September 2016 till March 2019. Delhi lies in the Northern parts of the Indian Subcontinent with 3 specific seasons: summer(March to June), monsoon(July –September), and winter(October –February). Winter seasons starts from October and lasts till February. In the present study, 3 winter seasons were considered. The distribution of various influenza and non-influenza viruses in different seasons is depicted in Figure 2.

**Figure 2.**
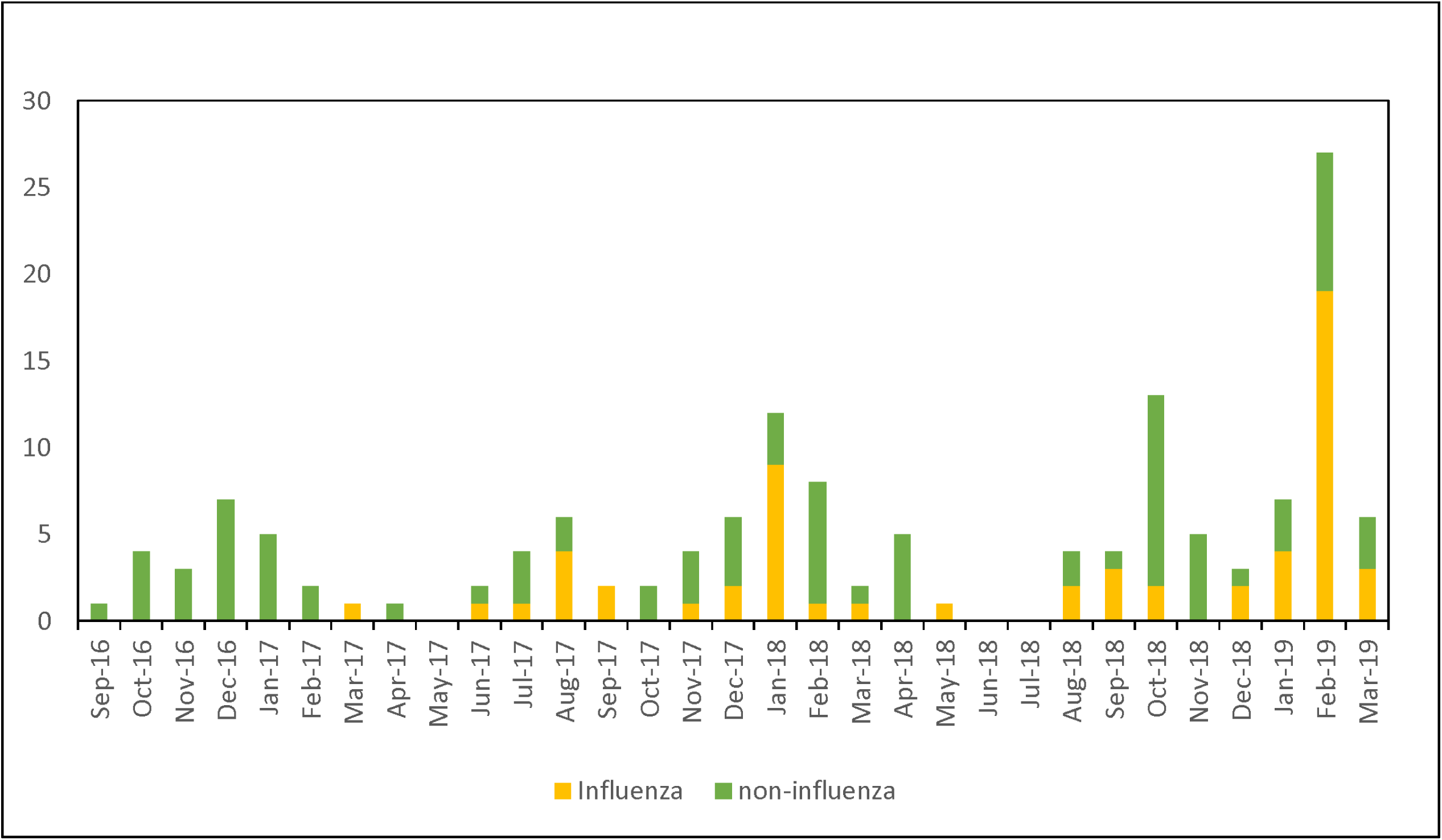
Seasonality of respiratory viral infections

Total number of Influenza and non-influenza cases seen in March-September were 62, 42.1% (IVI: 24, NIVI:38) lesser than 85 (57.8%) (IVI :39,NIVI:46) seen in October-February. Two peaks could be clearly seen first in the months of January and February and the other one in August and October.

### RVI Co-infections with other pathogens

Out of 147 patients, in 87 patients investigations for a bacterial and fungal etiology that was done at baseline were available (Figure 2).

In 44 (50.5%) patients also showed bacterial/fungal co-infection along with RVI. 32 (23.8%) patients with bacterial co-infection, 12 (37.5%) with fungal and 3 (6.3%) with both bacterial and fungal (3, 6.3%) was seen. Co-infections were maximum with Influenza group of viruses (19, 40.4%) followed by Rhino/entero group of viruses (12, 25.5%) among these 47 co-infected patients.

Development of subsequent/ secondary bacterial infection was also observed in these 147 patients. Secondary bacterial infection was considered when lower respiratory tract sample (sputum, BAL) became positive for a bacterial isolate after RVI detection up to a week in the patients negative for the respective bacterial infection initially. Overall secondary bacterial co-infections were seen in 15 (10.2%) patients which were mostly seen after Influenza viral infections 6 (40%) and Rhino viral infections 6 (40%) as compared to other RVI etiology.

### Association of respiratory viruses and mortality

Overall 67 deaths were observed in the RVI positive patients. Mortality was higher in NIVI group (43, 49.4%) as compared to IVI (24, 40%) [p=0.015] being maximum in Rhinovirus, 22 (32.8%) as shown in Table 4.

**Table 4:** Viral etiology wise mortality in the RVI cases

| **Respiratory viruses** | **Mortality in patients [n =67]**<br>**n (%)** |
| --- | --- |
| Influenza A (H1N1) pdm 2009 | 13 (19.4%) |
| Influenza A (H3N2) | 7 (10.4%) |
| Influenza B | 4 (5.9%) |
| RSV | 9 (13.4%) |
| MPV | 4 (5.9%) |
| PIV 1 | 1 (1.4%) |
| PIV 3 | 1 (1.4%) |
| Rhinovirus | 22 (32.8%) |
| Adenovirus | 2 (2.9%) |
| Coronavirus | 4 (5.9%) |

On comparing the RVI cases that died(n=67) with those who survived(n=80) a significant association was seen in the univariate analysis with the age of the patient (p=0.03), more seen in the age group 46-60 years of age, all the deaths occurred in the patients who were admitted and required the ICU care (p<0.001), with the underlying cirrhosis of the liver (p<0.001), bilirubin level (p<0.001), creatinine levels (p<0.001)and platelet count (p<0.001) [Table 5]. However on multivariate analysis, no association of outcome in terms of mortality was found with analyzed demographic and biochemical parameters.

**Table 5:** Association of mortality in RVI positive cases with various clinical and laboratory parameters.

| S. No | Variable | Mortality(n=67) | No Mortality(n=80) | p-value |
| --- | --- | --- | --- | --- |
| **1.** | **Age (in years)** |  |  | 0.03 |
|  | 18-30 | 4 (5.9%) | 22 (27.5%) |  |
|  | 31-45 | 19 (28.3%) | 28 (35%) |  |
|  | 46-60 | 27 (40.2%) | 23 (28.7%) |  |
|  | >60 | 17 (25.3%) | 7(8.7%) |  |
| **2.** | **Sex** |  |  | 0.25 |
|  | Male | 53 (79.1%) | 65 (81.2%) |  |
|  | Female | 14 (20.8%) | 15 (18.7%) |  |
| **3.** | **I/W/O** |  |  | 0.00 |
|  | ICU | 67 (100%) | 34 (42.5%) |  |
|  | Ward | 0 (0%) | 36 (45%) |  |
|  | OPD | 0 (0%) | 10(12.5%) |  |
| **4.** | **Cirrhosis** |  |  | 0.00 |
|  | Present | 60 (89.5%) | 39 (48.7%) |  |
|  | Absent | 7 (10.4%) | 41 (51.2%) |  |
| **6** | **Total Bilirubin** | 12.91±10.78 | 4.73±7.53 | 0.00 |
| **7.** | **AST** | 182.49±322.66 | 395.27±2238.65 | 0.00 |
| **8.** | **ALT** | 76.07±103.61 | 160.61±666.38 | 0.05 |
| **9.** | **ALP** | 111.21±107.64 | 116.47±75.89 | 0.40 |
| **10.** | **GGT** | 88.70±148.03 | 56.61±69.03 | 0.95 |
| **11.** | **Urea** | 95.17±56.15 | 44.51±32.23 | 0.04 |
| **12.** | **Creatinine** | 1.80±1.46 | 1.03±1.01 | 0.00 |
| **13.** | **Hemoglobin** | 8.01±1.74 | 9.78±2.62 | 0.73 |
| **14.** | **Platelet count** | 70.18±47.34 | 143.86±118.99 | 0.06 |

## DISCUSSION

To our knowledge, the present study is the first descriptive study on various respiratory viruses in adult population with liver disease. Influenza and non-influenza viruses have also been compared in terms of their clinical presentation and clinical outcome. Seasonality of the occurrence of these RVIs is also studied.

The present study highlighted that occurrence of RVI is more commonly seen in patients who required intensive care treatment, with higher Child Pugh score and more with cirrhosis. RVI occurrence was higher in CHILD category C as compared to lower categories. Overall prevalence seen in our study of RVI in liver disease patients was 24.3% which is similar to earlier studies from our institute where overall prevalence of RVIs was found in 22.2% cirrhotic patients with pneumonia [16].

Apart from IV other NIV should also be studied especially in patients requiring ICU care as NIV are more common in them. Earlier these groups of viruses were not considered to be causative for pneumonia but recent literature cites etiological role of rhino virus group in the causation of pneumonia in adults [3, 17]. In the present study rhino viruses were also associated with mortality as seen in 16 cases, though all the 16 cases were from ICU and multiple factors contribute to mortality. RVI in isolation cannot be contributory in mortality. Furthermore SARS-CoV-2 has established that NIV are an important cause of viral pneumonia and has necessitated the diagnosis of RVIs at the earliest.

Influenza A viruses were associated with more adverse outcome has been shown by earlier publications [18]. Influenza A is the greatest cause of mortality and morbidity among the viral types of pneumonia [19]. In our study influenza group of viruses accounted to 40.8% of the total RVIs. Few studies done in our center have shown the prevalence of influenza virus and its impact on the clinical outcome of cirrhotic patients [16], [20]. Interestingly in the present study mortality associated with NIV (49.4%) was more than IVIs (40%). This highlights the importance of constant surveillanve of NIVs especially in the critically ill.

In India, multisite epidemiological and virological influenza surveillance established previously reported peak influenza activity to be associated with rainy season and a secondary peak in the winter months [21]. Annual Influenza vaccination is recommended in patients with underlying liver disease especially cirrhotic group. Both IV and NIV were seen peaking in Delhi during monsoon period in the months of August – October and then again from December to February during the winter hence in Delhi a pre monsoon influenza vaccination strategy must be established. Our findings are consistent with previously reported studies where a monsoon peak followed by minor peak in winter was observed in Delhi [21]. Mixed viral-bacterial infections may be associated with an increased risk of mortality [22]. RVI alters respiratory flora and making the host more susceptible to secondary bacterial infections. Secondary bacterial infections were also quite common with Rhino associated RVI [23]. In the present study also secondary bacterial infections were seen mostly in Rhino associated RVI (40%).

Viral infections predispose patients to secondary bacterial infections, which often have a more severe impact on patients with cirrhosis. As the host immune system are already compromised in patients with cirrhosis it is therefore imperative to identify ARI caused by viruses in them and where ever a definitive treatment is available to initiate so that secondary bacterial infection and prolonged stay of the patient can be prevented.

Cirrhotic patients, especially the higher risk groups, should be targeted for preventive measures such as influenza and pneumonia vaccination as well as early and effective outpatient management of those who have contracted ARI in order to prevent costly and high-risk hospital admissions.

Although the composition of the gastrointestinal microbiome is largely influenced by dietary patterns, respiratory viral infections could also contribute, along with other stress inducers such as broad-spectrum antibiotics exposure and chronic inflammation.

Using animal models of pulmonary infections by influenza and respiratory syncytial virus (RSV), multiple groups have shown that the gut microbiome is clearly impacted by respiratory viral infections, despite the lack of detectable respiratory virus in the gut [24–28].

Gut microbiome alterations influence the severity outcome in patients with cirrhosis hence all the more essential to early diagnosis and treat any respiratory viral infection.

Previous studies have established the role of multiplex PCR panels as more rapid and sensitive than routine culture and antigen detection methods [29-36]. Although multiplex panels are rapid and accurate, the high cost to the laboratory and patient may limit their use in resource-limited settings of developing nations.

Our study does have few limitations. Firstly, it is a single center laboratory based study where majority are known patients with liver ailments. Secondly The lack of a control population with no respiratory symptoms is another limitation that precludes any conclusion on the pathogenicity of NIRV. Despite these minor limitations our study adds significantly to the literature by contributing to better characterization of the burden of NIV in adult patients with liver cirrhosis.

## Data Availability

Readers may contact the authors for accessing data used in this study.

## Acknowledgement

The authors would like to thank Mr. Sandeep Singh for compiling the data and all the technical staff of the Department of Clinical Virology, Institute of Liver and Biliary Sciences, New Delhi

## CONCLUSION

With the on going COVID-19 pandemic the role of NIV as a major cause of viral pneumonia has come into lime light. The otherwise neglected NIV shoud be evaluated and studied in more details to understand their pathophysiology as a cause of RVI in cirrhotic patients.

## DATA AVAILABILITY STATEMENT

Readers may contact the authors for accessing data used in this study.

## AUTHORSHIP CONTRIBUTION STATEMENT

**EG :** Writing - original draft, Writing - review & editing, Methodology, Supervision, Project administration. **AP -** Writing - review & editing **KA**: Resources, Methodology, Data analysis. **KR, RA**,**SSRP** : Data curation. **DB, RM:** Writing - review & editing, Supervision. **SKS :** Project administration, Supervision, Writing - review & editing.

